# One algorithm, tenfold range: measurement non-equivalence makes cross-national dementia prevalence unidentifiable

**DOI:** 10.64898/2026.11.20.26368574

**Authors:** Sumei Zhu, Yukai Luo

## Abstract

**Background:** Cross-national estimates of dementia prevalence rest on brief cognitive batteries scored against fixed cut-offs. Neither whether they measure the same construct everywhere nor how much of any cross-national difference is measurement has been formally quantified.

**Methods:** In 66,890 adults aged ≥65 from four population-based cohorts (USA 2016, England 2021–2023, 19 European countries 2021, China 2018) we applied a single harmonised algorithm (Langa–Weir, 27 points) and quantified how far the resulting prevalence moved across 1,152 specifications, measurement-invariance and equating models, latent class analysis, bias correction against a US gold standard, mortality and burden-of-disease anchoring, and multiple imputation.

**Results:** Weighted prevalence ranged from 1.25% (95% CI 0.82–1.83) in England to 12.79% (11.69–13.96) in China (10.2-fold; China minus USA 7.43 points, 6.22–8.65), but the median across specifications was 2.3-fold (IQR 1.5–4.4), the 10.2-fold value sitting at the 94.6th percentile. Scalar invariance was rejected (ΔCFI −0.066 vs metric), driven by serial-7 subtraction (odds ratios 2.9–3.4). No operating characteristics can reproduce both the US gold standard (10.0%) and the Chinese national estimate (6.0%): the identity implies Se+Sp−1 = −1.86, outside the admissible range, which would require Chinese prevalence of at least 23.9%. A published harmonised-protocol algorithm applied to the English HCAP subsample gave 7.1% (5.7–8.7), 5.7 times the brief-algorithm value in the same population; harmonised-protocol estimates span 1.4-fold rather than 4.3-fold.

**Conclusions:** Absolute cross-national dementia prevalence is not identifiable from brief screening algorithms. Rank order is more stable than level but is itself specification-dependent (China highest in 45%, England lowest in 65%), whereas the relative risk attached to the label is similar across cohorts (pooled mortality hazard ratio 2.14, I² = 0). We propose a seven-item minimum reporting standard, three items of which can be adopted immediately, and show that metric choice moves the global case count tenfold.

## Introduction

Dementia is among the largest contributors to global disability, and the numbers that frame policy are derived, not counted. The Global Burden of Disease (GBD) study estimated 57.4 million people living with dementia in 2019, rising to 152.8 million by 2050, with an age-standardised prevalence that barely changes (+0.1%) — a projection driven almost entirely by population ageing [1]. The 2021 iteration placed the global total at 56.9 million and reported an age-standardised rate of 694.0 per 100,000, with a range across the 204 modelled countries of 377.6 to 900.8 [2]. National and international targets, including the WHO Global Action Plan’s treatment targets, are expressed as diagnosis rates whose denominator is an "expected" case count [3].

In countries without clinical registries, that denominator comes from population surveys. Because full clinical assessment is too costly to field at scale, surveys derive dementia status algorithmically from brief cognitive batteries. The most widely used is the Langa–Weir algorithm, which applies education-adjusted cut-offs to a 27-point battery of immediate and delayed word recall, serial-7 subtraction and backward counting [4,5]. It underpins the US Health and Retirement Study (HRS), the English Longitudinal Study of Ageing (ELSA), the Survey of Health, Ageing and Retirement in Europe (SHARE) and the China Health and Retirement Longitudinal Study (CHARLS), and is the instrument through which cross-national comparisons are routinely made [6–8].

Whether a single algorithm measures the same construct across languages, education systems and survey modes has, however, rarely been tested formally. Three lines of evidence suggest it may not. First, item-level analyses of harmonised cognitive batteries show that individual items discriminate very differently across countries [9]. Second, unsupervised clustering of the same batteries yields prevalence estimates that differ from cut-off-based ones by a factor of two or more [10,11]. Third, in countries where a clinical gold standard exists — through the Harmonized Cognitive Assessment Protocol (HCAP), a standardised clinical assessment embedded in a subsample of each survey — the agreement between the algorithmic label and the clinical diagnosis is moderate at best [12,13]. What has not been established is the magnitude of the resulting uncertainty: whether the observed cross-national range reflects disease, measurement or some mixture of the two, and whether any set of operating characteristics could reconcile the observations at all.

We therefore asked four questions of four population-representative cohorts spanning the USA, England, Europe and China (251,263 person-wave records; 66,890 adults aged ≥65 contributing to prevalence estimation). First, what is the observed cross-national range under one identical algorithm? Second, how much of that range is an artefact of analytical choice, quantified across 1,152 defensible specifications? Third, is the algorithm measurement-invariant, and what happens to the range when the cohorts are placed on a common latent scale? Fourth, can any single set of operating characteristics reproduce both the observed values and the external gold standards — and if not, what can still be said about dementia burden across countries?

## Methods

### Study populations

In this cross-sectional analysis we used the most recent wave of each cohort for which the full 27-point battery and post-stratification weights were simultaneously available, with a proxy-informant module additionally present in HRS, ELSA and SHARE: HRS wave 13 (2016),

ELSA wave 10 (2021–2023), SHARE wave 9 (2021; 19 countries) and CHARLS wave 4 (2018). In total 66,890 respondents aged ≥65 contributed to weighted prevalence estimation (HRS 10,117; ELSA 3,713; SHARE 45,401; CHARLS 7,659; Table 1 and S1 Table). All four surveys are publicly available, use multistage probability sampling of community-dwelling adults and provide sampling weights. All analyses are weighted unless stated. Respondents who were interviewed and classified but whose survey weight was zero or missing (286 in HRS, 332 in ELSA, 3 in SHARE and 221 in CHARLS; 10.3% of classified ELSA respondents) were excluded from weighted estimation, so the estimating samples are slightly smaller than the classified samples shown in Table 1. The ELSA age variable is about three years behind the wave-10 interview; a sensitivity analysis using the wave-10 age variable is reported in S14 Table. Variances were estimated by Taylor-series linearisation using the sampling weights; because stratum and cluster identifiers are not retained for all four cohorts in the released files, clustering is not modelled and the intervals may therefore understate design-based uncertainty. The contrasts that carry the argument are between point estimates differing several-fold, far beyond any plausible correction of that kind. The pooled panel comprised 251,263 person-wave records. Sample sizes were determined by the size of each released cohort file and by the availability of the 27-point battery, not by an a priori power calculation; the smallest analytic sample (ELSA, 3,713) still gives a 95% confidence interval of about ±0.5 percentage points around a prevalence of 1.25%. Reasons for non-interview could not be disaggregated, because the released files carry no codebook for the response-status codes; the cascade from interview to assigned label is shown in S2 Fig.

**Table 1.** Cohorts, waves and analytic samples. Demographic composition of the analytic samples, adults aged ≥65 interviewed in the analysed wave.

| **Cohort** | **Country/region** | **Wave (year)** | **Interviews aged ≥65 (n)** | **Proxy module** | **Self-reported diagnosis** | **Death status** |
|---|---|---|---|---|---|---|
| HRS | USA | 13 (2016) | 10,117 | Yes (9-point) | Yes | Yes |
| ELSA | England | 10 (2021–23) | 3,713 | Partial (no memory rating) | Yes | No (waves 7–10) |
| SHARE | 19 European countries | 9 (2021) | 45,401 | Yes (34.4% with memory rating) | Yes | Yes |
| CHARLS | China | 4 (2018) | 7,659 | No | Yes | Yes |
| **Characteristic** | **HRS (USA)** | **ELSA (England)** | **SHARE (Europe)** | **CHARLS (China)** |  |  |
| N (interviewed, aged ≥65) | 10,117 | 3,713 | 45,401 | 7,659 |  |  |
| Age 65–74 | 4,451 (58.3%) | 2,423 (62.2%) | 25,022 (51.0%) | 5,141 (65.5%) |  |  |
| Age 75–84 | 4,114 (29.3%) | 1,105 (30.3%) | 15,649 (35.8%) | 2,096 (28.1%) |  |  |
| Age 85+ | 1,552 (12.5%) | 185 (7.5%) | 4,730 (13.1%) | 422 (6.4%) |  |  |
| Female | 6,014 (55.6%) | 1,974 (54.1%) | 25,720 (57.2%) | 3,944 (51.7%) |  |  |
| Education, low | 2,164 (16.3%) | 799 (31.9%) | 16,861 (41.9%) | 7,117 (91.2%) |  |  |
| Education, middle | 4,041 (39.0%) | 1,817 (50.6%) | 18,266 (37.0%) | 416 (6.6%) |  |  |
| Education, high | 3,886 (44.7%) | 808 (17.6%) | 10,274 (21.1%) | 126 (2.1%) |  |  |
| Proxy interview | 721 (5.0%) | 144 (3.8%) | 1,347 (3.1%) | 0 (0.0%) |  |  |
| No usable 27-item score | 721 (5.0%) | 481 (14.3%) | 9,465 (20.7%) | 3,414 (42.8%) |  |  |

Cells are unweighted n with the survey-weighted percentage of the interviewed sample in parentheses (age and sex are weighted to the cohort's own target population). Education is the highest qualification in three harmonised bands, with percentages over respondents with non-missing education (missing for 26 HRS, 289 ELSA, 0 SHARE and 0 CHARLS respondents). Proxy interviews are those in which the respondent could not be interviewed directly; the proxy module was absent in CHARLS and lacked a memory-rating item in ELSA, so proxy respondents there cannot be classified. The last row counts respondents without a scorable 27-point score; it is smaller than the unclassified count in S7 Table for SHARE because proxy respondents can be classified through the proxy module without a 27-point score.

S2 Fig. traces the derivation of each analytic sample. Of those interviewed, a cognitive classification could be assigned to 99.9% (10,111), 87.0% (3,232), 78.9% (35,826) and 55.4% (4,245): the unclassified proportions are 0.06%, 13.0%, 21.1% and 44.6%, concentrated among respondents with incomplete item-level data in ELSA, among proxy respondents and respondents with incomplete item-level data in SHARE, and among respondents with incomplete item-level data in CHARLS. Missingness is therefore far from negligible in the two cohorts with the most extreme prevalence, and we address it in two ways: multiple imputation under a missing-at-random assumption (m = 20) and a delta-adjustment sensitivity analysis that assumes missing respondents differ systematically from observed ones (Results; S7 Table).

### Dementia algorithm

We implemented the Langa–Weir 27-point algorithm exactly: immediate recall (0–10), delayed recall (0–10), serial-7 subtraction (0–5) and backward counting (0–2), with scores of 0–6 classified as dementia and 7–11 as cognitive impairment without dementia (CIND) [4,5]; alternative algorithmic classifications of the same battery are known to yield materially different prevalence estimates [14]. In HRS, our constructed score correlated r = 0.994 with the official released variable (r13cog27), and person-level differences were 0–2 points. Because CHARLS does not administer backward counting, we imputed the modal value of 2 points, the value observed in 92.9% of HRS respondents; a sensitivity analysis excluding the item is reported.

Respondents who could not be directly interviewed were assessed with an informant module scored 0–9 (proxy memory rating 0–4, five instrumental activities of daily living 0–5). In HRS and SHARE this produced a direct classification. In SHARE, where the memory-rating item is administered to only 34.4% of proxies, the proxy branch was calibrated within age × sex × education strata against the self-report branch; sensitivity analyses varying proxy prevalence by ±50% are reported in S2 Table.

### Measurement invariance

We fitted multi-group confirmatory factor models to the four cognitive items (immediate recall, delayed recall, serial-7, backward counting), treating the two count items as continuous and, in a sensitivity analysis, as ordinal with a weighted least squares means- and variance-adjusted estimator (WLSMV). Invariance was assessed sequentially (configural, metric, then scalar), with ΔCFI ≤ 0.010 and ΔRMSEA ≤ 0.015 taken as evidence of invariance [15]. Differential item functioning (DIF) for the serial-7 item was quantified with a proportional-odds model that included total score and cohort, and with the Mantel–Haenszel procedure (≥5 vs <5). Latent mean differences were estimated under partial scalar invariance, with and without adjustment for age, sex and education (MIMIC). Full model fit, loadings and item parameters are reported in S3 Table.

### Equating and latent class analysis

To ask what the cross-national range would be on a common metric, we used the partial-scalar solution to place each cohort’s latent distribution on the HRS metric (mean-only and mean-plus-variance equating) and recomputed in each cohort the proportion falling below the HRS cut-off. As an alternative that requires no common metric, we fitted latent class models separately within each cohort (K = 3 selected by BIC; posterior-weighted class prevalence; 400 bootstrap replicates) under three indicator designs (S4 Table and S5 Table).

### Multiverse analysis

We enumerated seven analytical dimensions: outcome definition (Langa–Weir ≤6, ≤9, self-reported physician diagnosis, or a relative cut-off at each cohort’s weighted 10th percentile); weighting (weighted/unweighted); proxy handling (include calibrated proxy branch / self-report only); age restriction (≥65, 65–79, ≥80, age-standardised); education restriction (all, low, high); cohort set (four cohorts, three without CHARLS, HRS + SHARE only); and standardisation (crude/standardised). This yielded 1,152 specifications, for each of which we computed the maximum-to-minimum prevalence ratio across cohorts.

### Gold-standard anchoring and a formal impossibility test

Clinical dementia prevalence from standardised HCAP assessments came from published sources: HRS-HCAP 2016 (10.0%, 95% CI 9–11) [12], NICOLA-HCAP in Northern Ireland (6.2%, n = 1,037) [16], SHARE-HCAP (7.0%) [17], Mex-Cog (6.7%) [18] and LASI-DAD (7.2%) [19]. Because no peer-reviewed HCAP prevalence was available for England itself at the time of analysis (S10 Table documents sources and access routes), we derived one directly from the ELSA-HCAP wave 1 data (n = 1,273), implementing the published HCAP diagnostic algorithm as described for NICOLA-HCAP [16]: five cognitive domains scored by second-order confirmatory factor analysis, with domain scores rank-normalised and adjusted for age, sex and education to a T metric (mean 50, SD 10), domain impairment defined at T ≤ 35, and dementia defined as impairment in ≥ 2 domains together with informant confirmation (Jorm ≥ 3.4 or Blessed ≥ 2). The estimate is weighted (r1hwtresp) and is an algorithmic classification, not a clinical adjudication. We applied the same pipeline to Mex-Cog (n = 1,691), where a published harmonised-protocol prevalence of 6.7% exists, as an external check of the implementation. The Northern Ireland value (6.2%) [16], the HCAP-network preprint for England (7.5%) [20] and the ELSA-HCAP project report (13%) [21] were retained for external comparison. For China, no CHARLS-HCAP prevalence has been published; we therefore used the national door-to-door survey estimate of 6.0% (95% CI 5.8–6.3; n = 46,011, aged ≥60) [22].

Any screening algorithm satisfies P_obs = Se·P_true + (1−Sp)(1−P_true) = α·P_true + (1−Sp), where α = Se+Sp−1 must lie in [−1, 1]. Applying this identity to two populations with externally known true prevalence yields two equations in two unknowns. We solved them for α and compared the solution with the admissible range, and we computed, for every α consistent with the US anchor, the implied Chinese prevalence, and conversely the true Chinese prevalence required to reproduce the observed value. The complete grid over alternative anchors and specificities is given in S9 Table.

### Mortality, missing data, standardisation and global re-scaling

Mortality follow-up was available for HRS, SHARE and CHARLS (ELSA does not release death status for waves 7–10, the waves that carry the full battery); we fitted Cox models adjusted for age and sex, with cohort interaction tests and a DerSimonian–Laird pooled estimate. Missing cognitive data were handled with multiple imputation by chained equations (m = 20, predictive mean matching) and with a delta adjustment. Prevalence was directly standardised to the WHO world standard population, the 2013 European standard population and the HRS age structure (S11 Table). To translate measurement into burden, we re-scaled the GBD 2021 global estimate by the ratio of each cohort’s prevalence to its country’s GBD 65+ prevalence, and by the ratios implied by the HCAP comparisons.

### Software and reproducibility

Analyses were conducted in R 4.4.2 (lavaan, survey, mirt, mice, survival, ggplot2). Scripts, derived datasets and figure-generating code are archived at Zenodo (DOI: 10.5281/zenodo.24166170) and described in the Data availability statement. The study follows the STROBE reporting guidelines for cross-sectional studies.

## Results

### Observed cross-national range under one identical algorithm

Applying the same 27-point algorithm to all four cohorts produced a weighted dementia prevalence of 5.36% (95% CI 4.89–5.86) in the USA, 1.25% (0.82–1.83) in England, 5.26% (4.83–5.71) in 19 European countries, and 12.79% (11.69–13.96) in China (Table 2, Figure 1). The maximum-to-minimum ratio was 10.2. The absolute difference between China and the USA was 7.43 percentage points (95% CI 6.22–8.65), whereas the Europe–USA difference was −0.10 points (−0.74 to 0.55), indistinguishable from zero. England lay 4.11 points below the USA (−4.78 to −3.43). Timing cannot explain the spread: the two most nearly contemporaneous cohorts — England (2021–23) and the 19 European countries (2021) — differ by a factor of 4.21 (95% CI 2.84–6.23; 3.58, 2.34–5.47, after age standardisation), and the USA (2016) and China (2018) differ by 2.39 (2.11–2.70; S14 Table).

**Figure 1.**
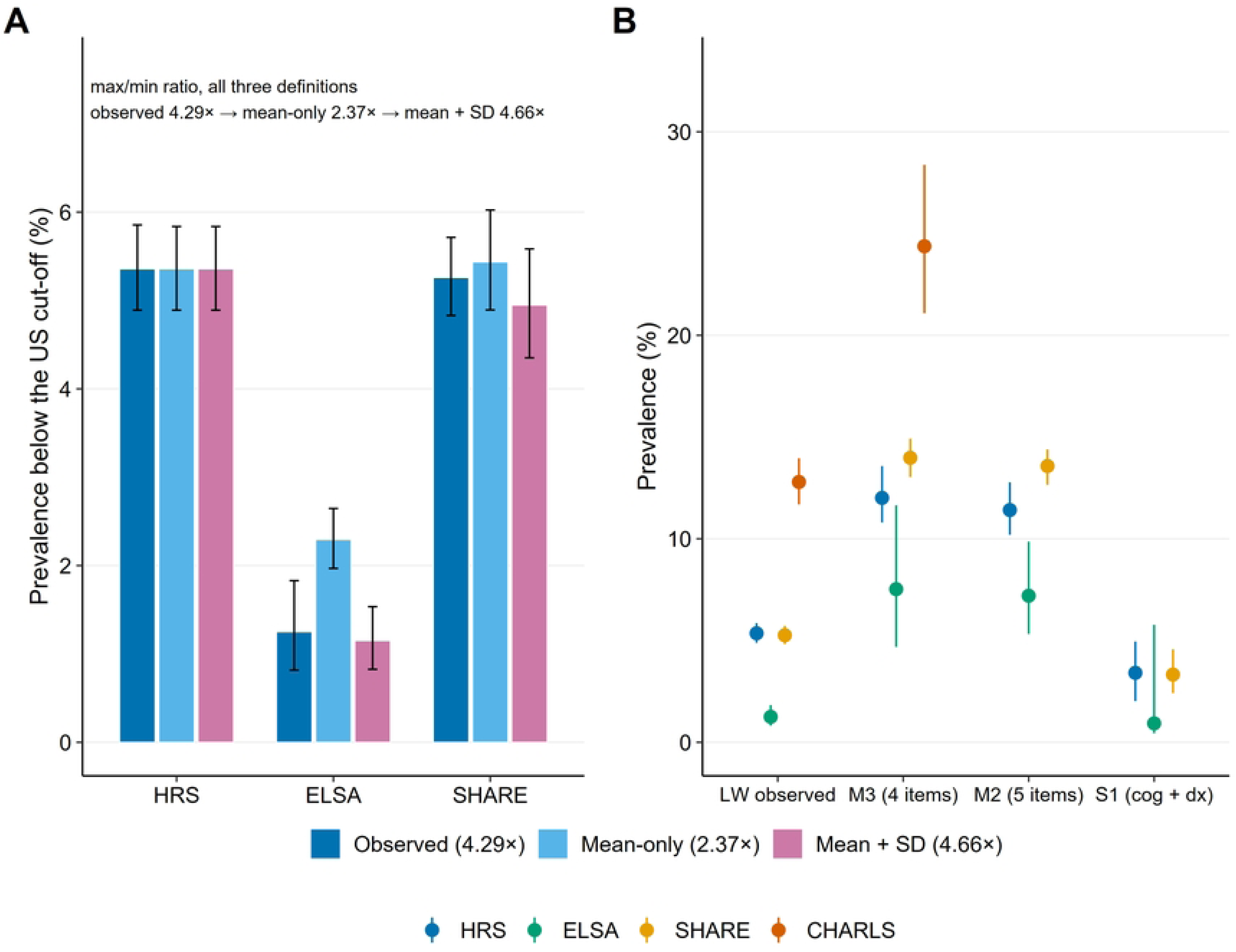
Prevalence of algorithm-defined dementia in four population-based cohorts. (A) Weighted prevalence of Langa–Weir ≤6/27 among adults aged ≥65, with 95% confidence intervals: USA 5.36% (4.89–5.86), England 1.25% (0.82–1.83), Europe 5.26% (4.83–5.71), China 12.79% (11.69–13.96); the four values span 10.2-fold. (B) The same prevalence stratified by sex. All estimates are survey-weighted; intervals are design-based (beta method). Dementia here and throughout is an algorithmic classification, not a clinical diagnosis.

**Table 2.** Weighted dementia prevalence (Langa–Weir ≤6/27), adults aged ≥65.

| **Cohort** | **Prevalence, % (95% CI)** | **Age-standardised (WHO), %** | **Diagnosis rate among cases, %** | **Diagnosis rate ratio (positive/negative)** |
|---|---|---|---|---|
| USA | 5.36 (4.89–5.86) | 5.44 (4.98–5.91) | 35.9 | 14.2 |
| England | 1.25 (0.82–1.83) | 1.45 (0.84–2.05) | 16.0† | 22.3† |
| Europe | 5.26 (4.83–5.71) | 5.17 (4.75–5.59) | 15.9 | 7.0 |
| China | 12.79 (11.69–13.96) | 16.31 (14.22–18.40) | 8.7 | 1.6 |

†Only 34 algorithm-positive respondents in ELSA; the diagnosis rate and its ratio are shown for completeness and should be interpreted with caution. The diagnosis rate is the weighted percentage reporting a physician diagnosis among all classified cases (self-reported and calibrated proxy cases); the ratio divides this rate by the diagnosis rate among algorithm-negative respondents, so that both columns use the same case definition.

Direct standardisation to the WHO world standard population left the ordering unchanged and widened rather than narrowed the China–USA gap (5.44, 1.45, 5.17 and 16.31%; gap 10.87 points), because the Chinese analytic sample is younger than the American (65.5% aged 65–74 and 6.4% aged ≥85, against 58.3% and 12.5%; Table 1). Women had a higher prevalence than men in every cohort (odds ratios for men 0.62–0.77), with no evidence of cohort heterogeneity in that pattern (interaction p = 0.40; Q = 2.93, I² = 0).

Removing backward counting altogether and re-cutting the three-item scale at the equivalent threshold (≤4 of 25) left the cross-national ratio essentially unchanged (11.4 versus 10.2), so the result is not an artefact of the Chinese imputation. Neither is it an artefact of the unclassified: even if all 3,414 unclassified CHARLS respondents were dementia-free, Chinese prevalence would be 7.31%, still the highest of the four cohorts and still a 5.9-fold range (S14 Table). The Chinese value is, however, unchanged by construction, and its placement on the 27-point scale is arbitrary: assigning 0 rather than the modal 2 imputed points would raise it to 22.3% (S13 Table). Among people classified as having dementia, the proportion reporting a physician diagnosis also varied by more than fourfold: 35.9% in the USA, 16.0% in England, 15.9% in Europe and 8.7% in China. The ratio of diagnosis rates between algorithm-positive and algorithm-negative respondents was 14.2, 22.3, 7.0 and 1.6 respectively: the same cut-off aligns with diagnosed dementia in England but barely at all in China.

### How much of the range is analytical choice?

Across 1,152 defensible specifications (S6 Table; 7 dimensions crossed in full), the maximum-to-minimum prevalence ratio had a median of 2.29 (interquartile range 1.53–4.41; full range 1.00–43.40) (Figure 2). The pre-specified baseline specification — Langa–Weir ≤6, weighted, with the calibrated proxy branch, all adults ≥65, all education levels, all four cohorts — yielded the 10.2-fold result, which sits at the 94.6th percentile of the 1,088 specifications with an estimable ratio (1,029 specifications at or below the baseline ratio, two of them tied with it, and 1,027 strictly below). Only 21.2% of specifications reached a ratio above 5, and 6.2% above 10; 44.7% did not reach even 2-fold. The composition of the specification space matters: among the 320 estimable specifications that include all four cohorts (384 in total) the median ratio was 4.66 (IQR 2.98–8.18), with 46% above 5-fold and 17% above 10-fold, whereas the 384 two-cohort specifications (USA and Europe only) had a median of 1.52 and none exceeded 5-fold (S14 Table).

**Figure 2.**
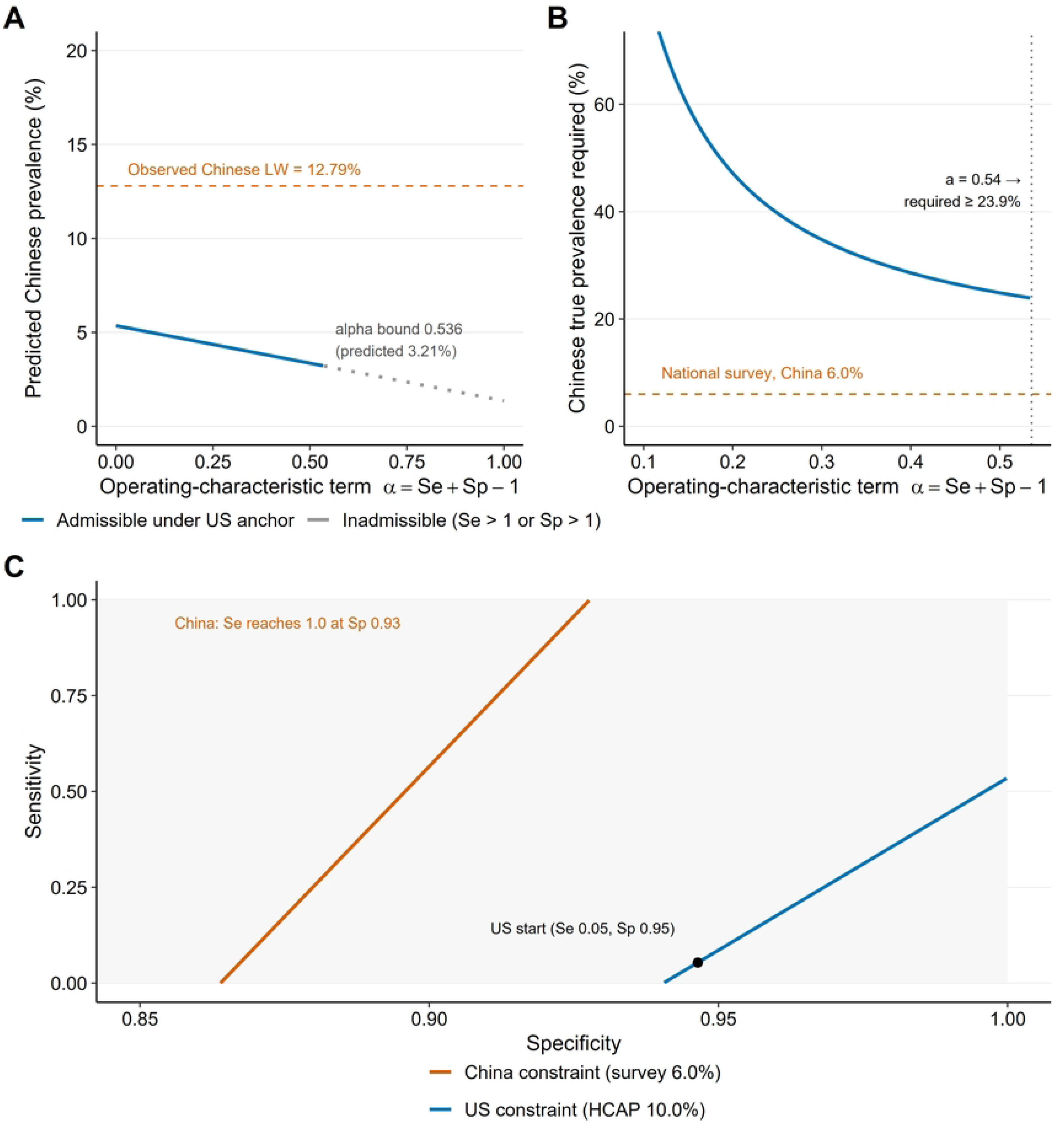
Specification curve across 1,152 defensible analytical specifications. (A) Specifications ranked by their estimated maximum/minimum prevalence ratio, on a logarithmic scale. The shaded band spans the interquartile range (1.53–4.41) and the dashed line marks the median (2.29); the orange point marks the pre-specified baseline analysis (ratio 10.23), which sits at the 94.6th percentile of the 1,088 specifications with an estimable ratio. (B) The same ranking decomposed by analytical dimension (outcome definition, weighting, proxy-branch handling, age restriction, education harmonisation, cohort set and standardisation), with the colour of each cell identifying the level of that dimension.

Rankings were likewise specification-dependent: China was the highest-prevalence cohort in 44.8% of four-cohort specifications and England the lowest in 64.8%, with both ordering claims holding simultaneously in 41.9% (50.3% of the 320 specifications in which the comparison is defined). The largest analytical levers were not weighting, proxy handling, age restriction or education (each with median ratios of 2.1–2.6), but the cohort set itself (median 4.66 for all four cohorts versus 1.52 for HRS and SHARE alone), and the choice between an absolute and a relative cut-off (4.36 for Langa–Weir ≤6 versus 1.52 for a cohort-specific 10th-percentile threshold).

### The algorithm is not measurement-invariant

Fitting multi-group confirmatory factor models to the four cognitive items (n = 48,011 across HRS, ELSA and SHARE) showed that configural structure was retained (CFI 0.949, RMSEA 0.142) but scalar invariance was rejected: ΔCFI −0.066 relative to the metric model and ΔRMSEA +0.023 relative to the metric model (Figure 3). Metric invariance was supported at our pre-specified threshold (ΔCFI −0.002), but was rejected once group sizes were balanced (ΔCFI −0.032), which suggests that loadings differ between cohorts as well as intercepts. Allowing the serial-7 intercept to vary restored most of the fit (ΔCFI +0.045), and the resulting partial-scalar model was retained.

**Figure 3.**
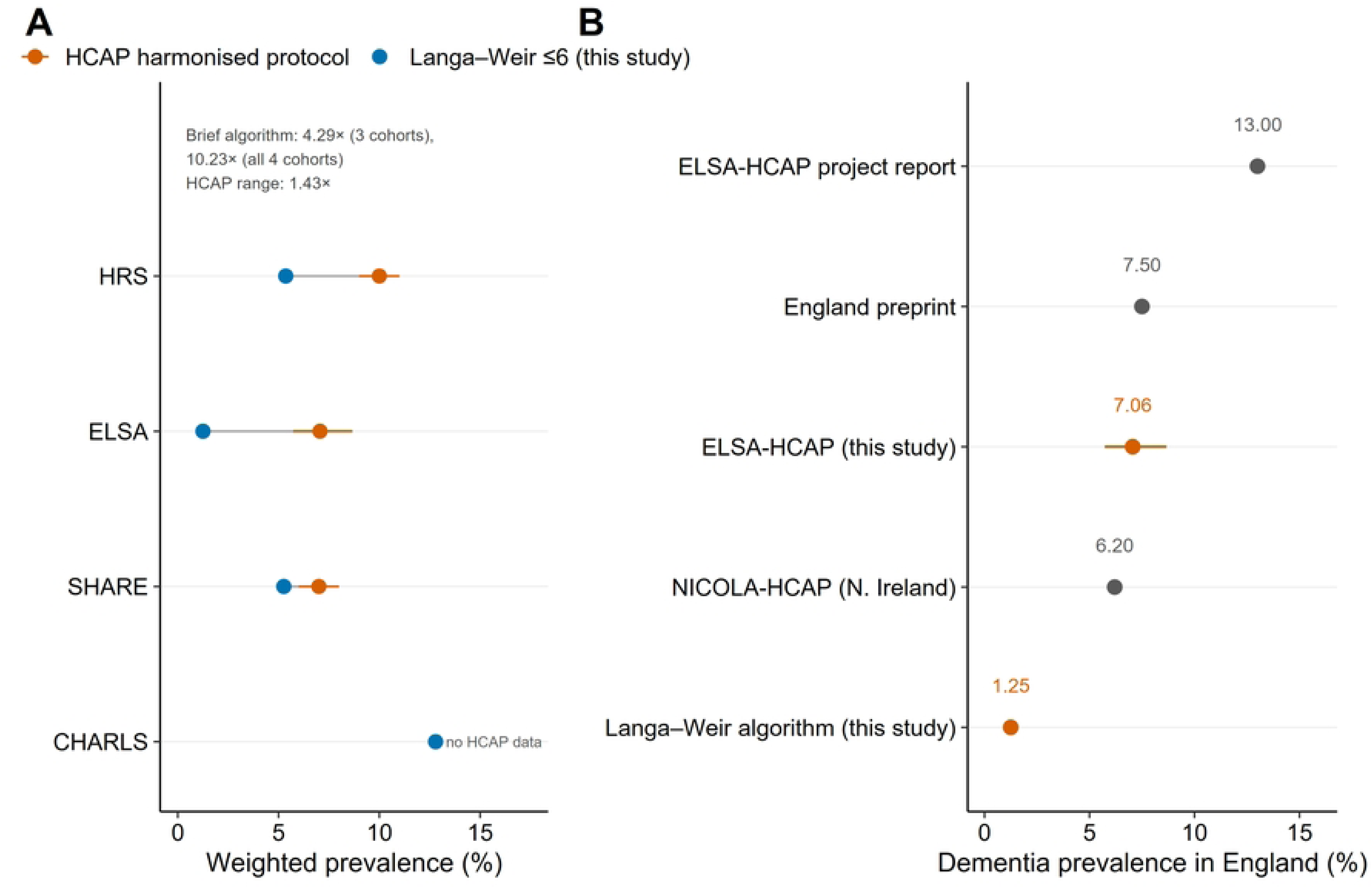
The brief algorithm is not measurement-invariant across cohorts. (A) Comparative fit index for configural, metric, scalar and partial-scalar models, annotated with the change in CFI relative to the configural model; scalar invariance is rejected (CFI 0.069 below the configural model, 0.066 below the metric model). (B) Unstandardised factor loadings by item and cohort. (C) Uniform differential item functioning odds ratios for serial-7 subtraction relative to the US cohort: ELSA 2.89 and SHARE 3.37 in the four-item three-cohort model, and CHARLS 1.68 in the three-item model that includes China. Serial-7 subtraction is the item that carries the invariance failure.

The item parameters identify the source. The serial-7 item loaded 0.48 in the USA, 0.17 in England and 0.34 in Europe, and its intercept in the partial-scalar model was 3.353 (USA), 4.175 (England) and 4.205 (Europe): at the same level of latent cognition, an English or European respondent scored 0.82–0.85 points higher. Backward counting carried almost no information in any cohort (loadings 0.02–0.09), so the 27-point score is not unidimensional. Differential item functioning for serial-7 was substantial in every pairwise comparison against the USA (proportional-odds odds ratios 2.89 for England, 3.37 for Europe, 1.68 for China; all p < 0.001), with the predicted probability of a perfect score at mean ability being 0.350 in the USA, 0.609 in England, 0.645 in Europe and 0.459 in China.

Under partial scalar invariance, latent mean differences relative to the USA were +0.386 SD for England, approximately zero for Europe, and −1.13 SD for China. In a MIMIC model of the same items, adjustment for age, sex and education moved the English difference from +0.40 to +0.34 SD, the contributions of age, education and sex being −0.143, +0.067 and +0.021 SD and reduced the Chinese difference by 43% (−1.13 to −0.65 SD); across the three specifications the adjusted Chinese difference ranges from −0.23 to −0.89 SD. Item-level difficulty therefore accounts for most of the divergence, with age, sex and education accounting for a substantial minority of it (43% of the Chinese difference and a much smaller share of the English one).

### Equating and operating characteristics cannot reconcile the cohorts

Placing the cohorts on a common latent metric (Figure 4) reduced the spread among the three high-income cohorts from 4.29-fold to 2.37-fold (mean-only equating) and 2.13-fold after covariate adjustment — roughly halving it. It did not reconcile China: with the three-item model required for a four-cohort comparison, the spread widened to 15.3-fold (mean-only) and 26.3-fold (mean-plus-variance), and even the most conservative adjusted estimate remained 6.9-fold. Under equating, the Chinese prevalence would be 31.8% (unadjusted) or 17.0–17.9% (adjusted) rather than the observed 12.8%: once the metric is held constant, the brief algorithm appears to under-rather than over-estimate Chinese prevalence. The mechanism is distributional: the Chinese immediate-recall mean is 2.92 of 10 against 5.00 in the USA, so the latent normal left tail sits proportionally about 2.5 times further below a sum-score threshold.

**Figure 4.**
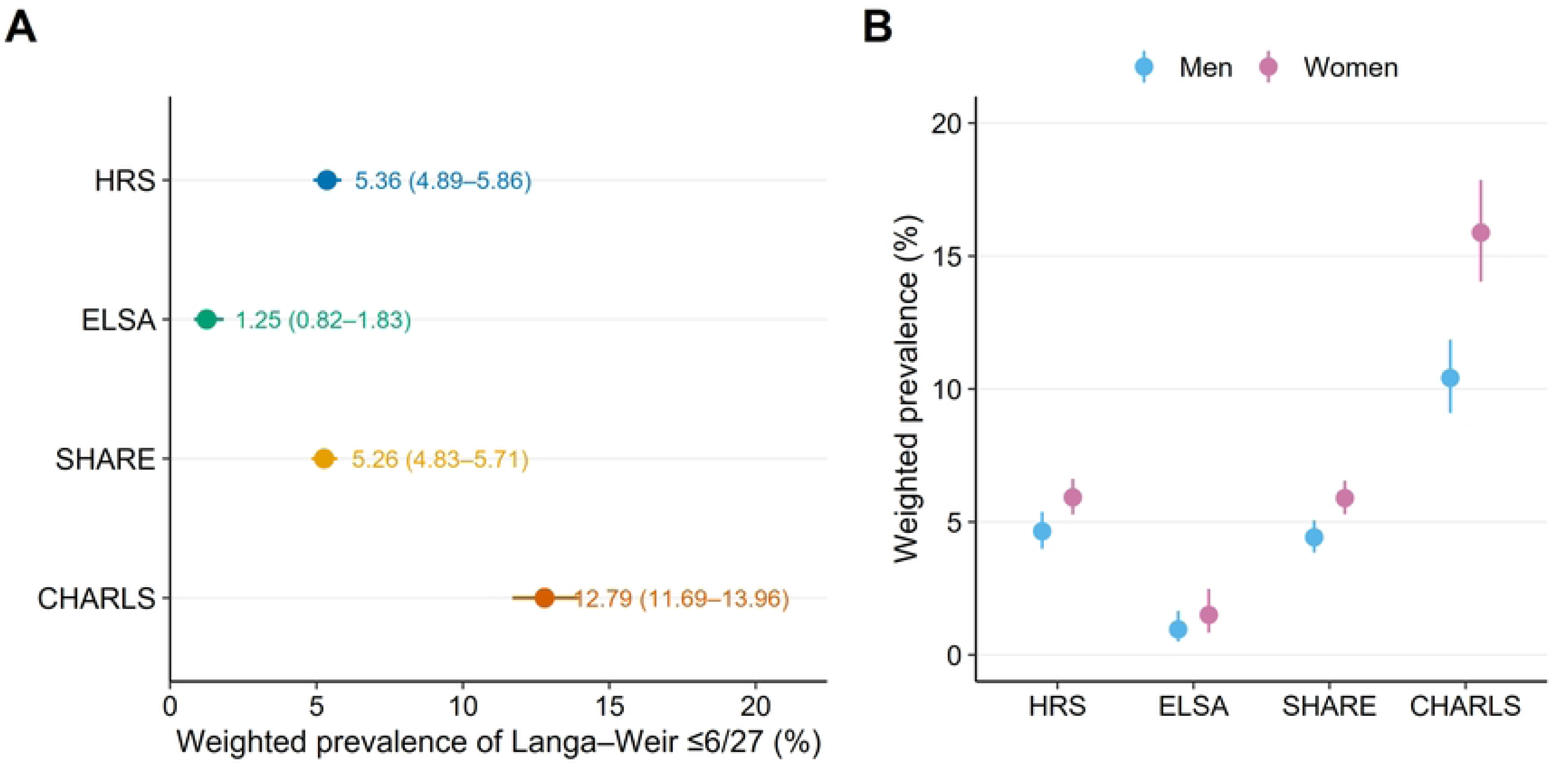
Equating and latent class analysis do not reconcile the cohorts. (A) Prevalence below the US latent cut-off under the observed score, mean-only equating and mean-plus-variance equating, for the three cohorts that administer the four-item battery; their maximum/minimum ratio is 4.29-fold under the observed score, 2.37-fold under mean-only equating, 4.66-fold under mean-plus-variance equating and 2.13-fold under mean-only equating with covariate adjustment. China, which requires the three-item battery, cannot be placed on this equating solution; in the four-cohort model its inclusion moves the ratio in the opposite direction, from 10.2-fold to 15.3-fold (Table 3). (B) Latent class prevalence under three indicator designs (four-indicator 3.24-fold, five-indicator 1.89-fold, three-indicator 3.67-fold); the classes are fitted separately within each cohort, so these estimates are supportive rather than a common-scale measurement.

**Table 3.** The same four cohorts under nine alternative metrics.

| Metric | USA | England | Europe | China | Max/min |
|---|---|---|---|---|---|
| Langa–Weir ≤6 (primary) | 5.36 | 1.25 | 5.26 | 12.79 | 10.2 |
| Langa–Weir ≤9 | 13.18 | 4.96 | 13.05 | 27.65 | 5.6 |
| Self-reported physician diagnosis | 4.34 | 2.52 | 5.06 | 7.17 | 2.8 |
| Cognition ≤6 + functional limitation | 2.56 | 0.48 | 2.21 | 5.86 | 12.3 |
| Equated to latent metric (mean-only; three-item, four cohorts) | 5.36 | 2.08 | 4.84 | 31.8 | 15.3 |
| Latent class analysis (4-indicator) | 12.01 | 7.52 | 13.98 | 24.38 | 3.2 |
| Multiple imputation (m = 20) | 5.37 | 1.72 | 6.60 | 16.58 | 9.7 |
| HCAP harmonised protocol | 10.0 | 7.1† | 7.0 | — | 1.4 |
| Specification median (1,152) | — | — | — | — | 2.3 |

†England: algorithmic classification applied by us to the ELSA-HCAP wave 1 sample (7.06%, 95% CI 5.73–8.67; n = 1,050). Northern Ireland (NICOLA-HCAP, n = 1,037) gives 6.2%; the HCAP-network preprint gives 7.5% and the ELSA-HCAP project report 13%, which would move the range to 1.43 and 1.86 respectively. All England values are algorithmic classifications rather than clinical adjudications.

As a cross-check that requires no common metric, latent class models fitted separately within each cohort gave posterior-weighted "most impaired class" prevalence of 12.01%, 7.52%, 13.98% and 24.38% — a 3.24-fold spread (1.89- to 3.67-fold across indicator designs) and within-class sensitivity of the screening cut-off ranging from 0.150 to 0.459, indicating that the same threshold captures very different fractions of the impaired population (S5 Table). We treat these models as supporting evidence only: because each cohort is fitted separately, class boundaries follow each national distribution.

Using the US HCAP clinical assessment as an anchor (true prevalence 10.0%, 95% CI 9–11, in the same survey and year in which the algorithm returned 5.36%), the implied sensitivity of the algorithm was 0.446 at a specificity of 0.99 (0.401 at 0.985; 0.491 at 0.995). Applying that single pair to all cohorts gave corrected prevalence of 10.0% (USA, by construction), 0.57% (England), 9.78% (Europe) and 27.06% (China) — a spread of 16- to 47-fold, larger than the observed one (Figure 5). The English observation of 1.25% alone requires a specificity of at least 0.986 for the algorithm to be non-negative in true prevalence (0.9875 on the 0.0025 grid search).

**Figure 5.**
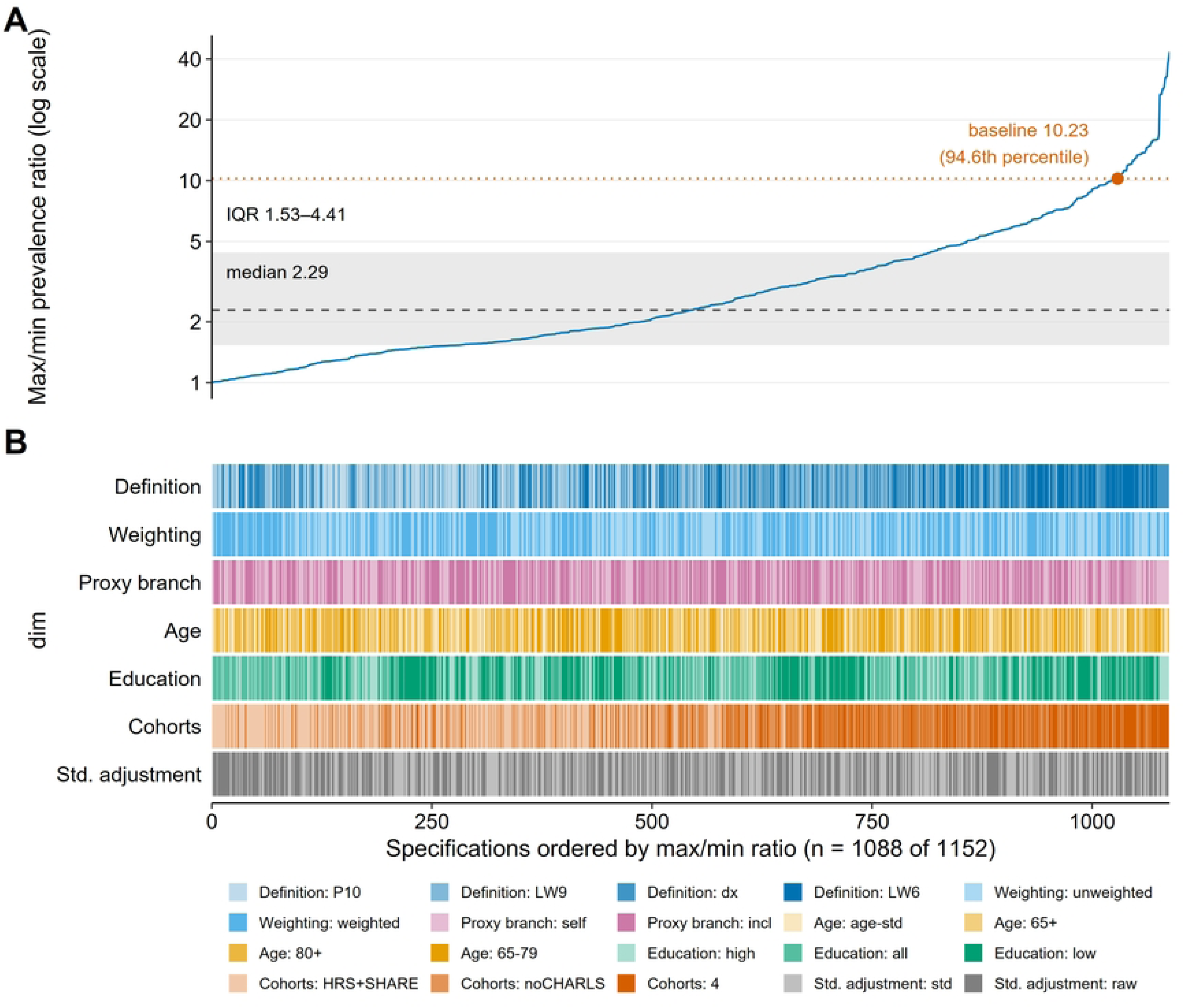
No pair of operating characteristics can reconcile the US anchor with the Chinese national survey. (A) Predicted Chinese prevalence as a function of the operating-characteristic term α = Se + Sp − 1, under the US harmonised-protocol anchor (10.0%); the observed Chinese value of 12.79% is never reached for any admissible α. (B) True Chinese prevalence required to reproduce 12.79% at each admissible α; at the upper admissible bound (α = 0.54) the required value is ≥23.9%, four times the national survey estimate of 6.0%. (C) Combinations of sensitivity and specificity satisfying the US constraint and the Chinese constraint; the curves do not intersect inside the unit square, and the Chinese constraint requires sensitivity above 1 for specificity ≥0.93.

The decisive test is algebraic. Any algorithm satisfies P_obs = α·P_true + (1−Sp) with α = Se+Sp−1 necessarily in [−1, 1]. Applying this identity to the USA (true 10.0%, observed 5.36%) and to China (true 6.0% from the national door-to-door survey, observed 12.79%) gives α = −1.86, outside the admissible range; equivalently, to reproduce the observed Chinese value under any operating characteristics consistent with the US anchor, Chinese true prevalence would have to be at least 23.9%, four times the national survey estimate. Across a grid of plausible true-prevalence values (USA 9–11%, China 5–8%) the implied α ranged from −1.24 to −7.43: there is no admissible solution in any cell (Figure 5).

### A harmonised protocol yields a 1.4-fold spread instead of 4.3-fold

Published HCAP estimates for the same or comparable populations (S10 Table) are 10.0% (USA), 6.2% (Northern Ireland), 7.0% (Europe), 6.7% (Mexico) and 7.2% (India) (Figure 6) [12,16–19]. Applying the published harmonised-protocol algorithm to the English HCAP subsample gave 7.06% (95% CI 5.73–8.67; MCI 24.45%, 21.37–27.82) among the 1,050 respondents with both cognitive and informant assessment — between the Northern Ireland value of 6.2% and the unpublished English figure of 7.5%. Across the three cohorts for which both quantities exist, harmonised-protocol prevalence spans 1.43-fold (7.0% to 10.0%), whereas the same brief algorithm spans 4.29-fold in those three cohorts (10.23-fold once China is added). The ratio of harmonised to brief-algorithm prevalence is 1.87 in the USA, 1.33 in Europe and 5.65 in England (5.65 with our own estimate; 4.96 with the Northern Ireland value, 6.02 with the 7.5% preprint and 10.4 with the 13% project report), which points to the British estimate as the most likely failure of the brief algorithm rather than a population with unusually low dementia burden. Equivalently, at most 17.7% of harmonised-protocol cases in England could have been captured by the brief algorithm even if every algorithmic case were a true case. Seven sensitivity specifications of the harmonised algorithm gave 5.86–8.63% once the cognitive-only specification was excluded (full range 5.86–14.21%). In the Mex-Cog validation sample the pipeline returned 4.04% (2.70–6.01) against a published value of 6.7%, and 5.15% (3.66–7.20) once the informant threshold was relaxed to compensate for the unavailability of four of the six Brief CSI-D items in that file — a partial validation, which we report as such.

**Figure 6.**
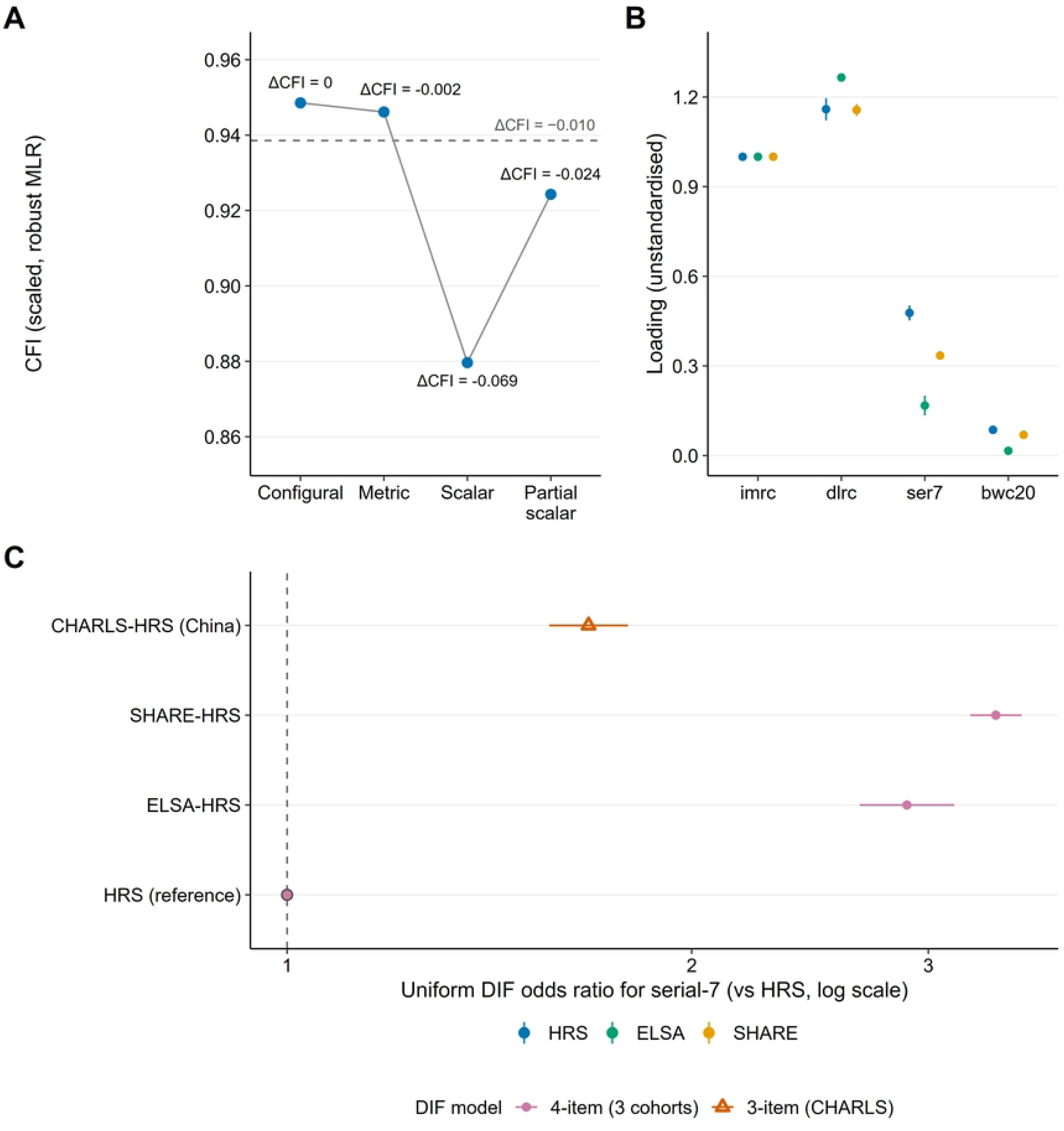
A standardised protocol narrows the three-cohort comparison from 4.3-fold to 1.4-fold. (A) Brief-algorithm prevalence (this study) against harmonised-protocol prevalence for the same or comparable populations: USA 5.36% vs 10.0%, Europe 5.26% vs 7.0%, England 1.25% vs 7.06%; no published Chinese harmonised-protocol prevalence exists. (B) Candidate English values: the estimate derived here from ELSA-HCAP wave 1 (7.06%, 95% CI 5.73–8.67), the Northern Ireland NICOLA-HCAP value (6.2%), the unpublished HCAP-network figure (7.5%) and the ELSA project report (13%). All English values are algorithmic classifications; whichever English value is used, the brief algorithm detects only a minority of harmonised-protocol cases.

### What survives: relative risk, ordering and the burden implied

Mortality follow-up (S8 Table; available for three cohorts; ELSA does not release death status in the waves carrying the full battery) showed an adjusted hazard ratio for algorithm-defined dementia of 2.20 (1.84–2.63) in the USA, 2.27 (1.86–2.77) in Europe and 1.83 (1.41–2.37) in China, with a pooled estimate of 2.14 (1.90–2.41). Heterogeneity was not detected by the pre-specified random-effects analysis (I² = 0.0%, p = 0.40), although label × cohort interaction tests ranged from p = 0.004 to p = 0.47 across specifications; the relative risk attached to the label is therefore broadly similar across cohorts even where its absolute meaning is not.

The absolute consequences of the label are not. Three-year excess mortality among algorithm-positive respondents was 97.5, 109.8 and 64.1 per 1,000 in the USA, Europe and China respectively, and the gradient in mortality across score bands was 13.1-fold, 10.1-fold and 3.0-fold; discrimination (C-statistic for the continuous score) was 0.675, 0.688 and 0.604. Multiple imputation for missing cognitive data moved every estimate upward and widened rather than narrowed the gap (5.37%, 1.72%, 6.60% and 16.58%; China/USA ratio 2.39 to 3.09), and the Chinese estimate remained above the US level even when every unclassified respondent was assumed dementia-free (7.31% versus 5.36%).

Re-scaling the GBD 2021 global estimate (56.9 million cases) by the ratios implied by our algorithm-based estimates yields between 9.4 million (if the English value were correct everywhere) and 79.9 million cases (if the Chinese value were) — a span an order of magnitude wider than the 95% uncertainty interval of the GBD estimate itself (±13%). Re-scaling by the HCAP-consistent ratios instead centres the estimate near the GBD figure, suggesting that the brief algorithm’s error, not the global model, is the dominant source of uncertainty in cross-national comparisons.

## Discussion

### Principal findings

One algorithm applied to four population-representative cohorts spanning the USA, England, Europe and China produced dementia prevalence estimates that differed 10.2-fold. That headline number is not, however, a fact about dementia. It is one draw from a distribution: across 1,152 defensible specifications the median ratio was 2.3, the interquartile range 1.5–4.4, and 45% of specifications did not reach even a two-fold difference. The algorithm failed formal tests of measurement invariance, and the failure is concentrated in a single item, serial-7 subtraction, which is scored differently in practice across surveys. Once the cohorts are placed on a common metric, the three high-income cohorts converge to a 2.1–2.4-fold range; China does not converge at all, and its estimated burden moves upward by between 33% and 148% depending on the equating model, although the direction of China’s deviation is not: every equating model that includes China moves it upward.

The most consequential finding is algebraic rather than empirical. No single pair of sensitivity and specificity can reproduce both the US gold-standard prevalence (10.0%) and the Chinese national survey estimate (6.0%) while also reproducing what the algorithm actually returns in each country: the implied value of Se+Sp−1 is −1.86, outside the interval [−1, 1] in which any screening test must live. Explaining the observed Chinese value would require a true Chinese prevalence at least four times that reported by the national survey. What differs between these populations, then, is more plausibly a property of the measurement than of the disease, and no recalibration within a single metric can repair it.

### Comparison with previous work

Our results extend three earlier observations into a quantitative statement. First, item-level analyses of harmonised cognitive batteries have shown that individual items discriminate differently across countries [9]; we show that this differential functioning is large enough to reject scalar invariance and, more importantly, to render the resulting prevalence ratio unidentifiable. Second, unsupervised approaches — clustering of harmonised batteries [10,11] and latent class models, as fitted here — produce estimates that differ substantially from cut-off-based ones, and our three-indicator designs span 1.89- to 3.67-fold ranges depending on design. Third, the recent European analysis of under-diagnosis in SHARE [23] documented that most people meeting algorithmic criteria lack a diagnosis; we find that the ratio of diagnosis rates between algorithm-positive and algorithm-negative respondents ranges from 22.3 in England to 1.6 in China, implying that the algorithm’s *meaning* — not merely its calibration — varies across settings.

A parallel literature has attempted to make these numbers comparable by other means. Unsupervised clustering of harmonised batteries produced global estimates several-fold above adjudicated ones in some settings [11]; retraining a clinical algorithm in a single reference sample [24] improves performance within that sample but does not establish that the retrained coefficients transport. Network-level work has instead emphasised protocol design and data-collection quality [25,26], and a construct-validity check using age gradients concluded that harmonised scores measure a broadly similar construct across six countries [27]. Our findings are consistent with that conclusion and locate the failure one level lower. Agreement on rank order and on the direction of age gradients [27] does not imply agreement on intercepts, and prevalence is an intercept quantity: it asks how many people fall below a fixed threshold, not whether they are ordered in the same way. Treating cross-cohort differences in that quantity as cohort effects, as recent generation comparisons have done [28], therefore risks attributing to ageing what belongs to the instrument.

The comparison with standardised clinical assessment is the most informative. Across the three populations in which both approaches can be applied (USA, Europe and England), harmonised-protocol prevalence lies between 7.0% and 10.0% — a 1.4-fold spread; across all five published harmonised-protocol estimates it spans 6.2% to 10.0%, or 1.6-fold [12,16–19]. Our brief algorithm, applied to the same populations, spans 4.3-fold, and 10.2-fold once China is included. The difference between the two is attributable to the measurement protocol, and it is larger than the real cross-national variation that GBD models across 204 countries (2.4-fold) [2]. The British case is the clearest: an algorithm-based estimate of 1.25% against a harmonised-protocol estimate of 7.06% that we derived from the same survey’s HCAP subsample, and against 6.2% in neighbouring Northern Ireland, 7.5% in an HCAP-network preprint and 13% in the project report — a five- to ten-fold discrepancy whichever value is used, in a country with a well-resourced survey.

### Interpretation: the label, the ordering and the risk

It is tempting to conclude that brief algorithms are simply invalid and should be abandoned. That would discard what they do carry. Relative risk is broadly portable: the mortality hazard ratio attached to the algorithmic label was 2.20, 2.27 and 1.83 across three countries, with a pooled estimate of 2.14; formal heterogeneity was absent under the pre-specified random-effects model (I² = 0, p = 0.40) though interaction tests ranged from p = 0.004 to p = 0.47 across specifications. Rank ordering is also more stable than level but not invariant: China is highest and England lowest in 42% of the 384 four-cohort specifications (50% of those with an estimable ratio), and the four structured analyses we report (equating, latent class, imputation, standardisation) each preserved that ordering. What is not portable is the absolute number, the absolute risk attached to it, and therefore any policy target expressed as a count of expected cases. These four cohorts are probability samples of community-dwelling adults aged ≥65 in their settings, so they exclude nursing-home residents and cannot be extrapolated to clinical or hospital populations, nor to low- and middle-income countries other than China.

This suggests a sharp distinction that the field has not made: prevalence as a *label* (how many people meet a criterion) and prevalence as a *quantity* (how many people have the disease) behave differently. Labels are cheap, reproducible within a cohort and useful for relative comparisons. Quantities are what GBD, WHO targets and national plans require, and they cannot be recovered from a label unless the label’s operating characteristics are known in that population — which is precisely what our impossibility result shows cannot be assumed.

### Implications for reporting and for global burden estimates

We propose a seven-item minimum reporting standard for cross-national prevalence work (Table 4), of which the first three are cheap enough to be adopted immediately; four of the seven are summarised here. (i) **Construct specification**: state which items, which cut-off, whether the cut-off is absolute or relative, and evidence of measurement invariance, or an explicit statement that the comparison assumes invariance. (ii) **Enrichment ratio**: report the ratio of diagnosis rates between algorithm-positive and algorithm-negative respondents — a minimal, cheap check of whether the label means the same thing in each setting (7.0–22.3 in the high-income cohorts versus 1.6 in China). (iii) **Specification range or equated interval**: report the median and interquartile range of the prevalence ratio across a pre-specified set of defensible specifications, rather than a single point estimate; and (iv) **anchoring bound**: the implied operating characteristics when an external gold standard exists, with an explicit statement when no admissible solution exists.

**Table 4.** Minimum reporting standard for cross-national dementia prevalence derived from brief algorithms (a checklist).

| # | Item | What must be reported | Worked example from this study | Consequence if omitted |
|---|---|---|---|---|
| 1 | Construct specification | Items administered, maximum score, absolute vs relative cut-off, education adjustment | 27 points (10+10+5+2); cut-off ≤6/27; education-adjusted | Readers cannot tell whether two estimates measure the same thing |
| 2 | Invariance evidence | Configural/metric/scalar fit (CFI, RMSEA) or an explicit no-invariance caveat | ΔCFI −0.066 (vs metric) for scalar; serial-7 intercept free | A 10-fold difference may be entirely metric |
| 3 | Enrichment ratio | Diagnosis rate in algorithm-positive versus algorithm-negative respondents | 14.2 / 22.3 / 7.0 / 1.6 across the four cohorts | Validity of the label in each setting is untested |
| 4 | Specification range | Median and IQR of the max/min ratio across pre-specified specifications | 2.29 (1.53–4.41) over 1,152 specifications | A single point estimate overstates precision by an order of magnitude |
| 5 | Equated interval | Prevalence after latent equating, with model fit | 2.1–2.4-fold among high-income cohorts; China/England 6.9-fold after age adjustment (15.3-fold unadjusted) | Direction of national deviation may be wrong |
| 6 | Anchoring bound | Operating characteristics anchored on ≥1 external gold standard, with the implied α | α = −1.86 (inadmissible) | An impossible calibration may pass unnoticed |
| 7 | Data and code | Released scoring code, item-level data or a reproduction pipeline | Scripts and derived datasets archived at Zenodo (DOI: 10.5281/zenodo.24166170); pipeline reproduces every table and figure | The estimate cannot be audited or updated |

†The checklist is ordered by the sequence in which a reader needs the information: what was measured (1–2), whether it means the same thing (3), how uncertain the number is (4–5), and whether it can be true at all (6). Items 1–3 are cheap and can be added to any existing survey report; items 4–6 require the analytical work we describe here.

The same logic applies to inequality. Educational gradients in the dementia label were steep and consistent in direction (low education 5.6–9.2 times high education, excluding China, where no high-education respondent met the cut-off; concentration indices −0.12 to −0.42), but their magnitude and their rank order across cohorts changed completely when the definition changed: under self-reported physician diagnosis the low–high gap fell to 1.3–4.9 percentage points in the high-income cohorts and reversed to −6.4 percentage points in China, and China moved from first to last (S12 Table, S1 Fig.). Reporting standards for inequality should therefore be as explicit as those for prevalence: which definition, which gradient metric, and whether the ordering is stable across definitions.

Applied to global burden, these standards change the headline. Re-scaling the GBD 2021 total by our algorithm-based estimates alone yields anything between roughly 9 and 80 million cases, a range an order of magnitude wider than the model’s own uncertainty interval. The HCAP-consistent ratios, by contrast, are close to unity, which localises the problem in the measurement instrument rather than in the global model. The policy consequence is not that the global estimate is wrong, but that its principal uncertainty is unmeasured and currently unacknowledged: a country whose survey uses a brief battery can move its apparent burden several-fold by changing an item’s scoring.

## Limitations

First, the English harmonised-protocol estimate is our own algorithmic classification of the ELSA-HCAP sample, not a clinical adjudication of the kind underlying the US figure (10.0%). It agrees with the unpublished English value (7.5%) and with Northern Ireland (6.2%) and is reproducible from the released data, but its absolute level could differ from an adjudicated value by several percentage points: in the Mex-Cog validation the same pipeline returned 4.0% against a published 6.7%, and 5.2% after an informant-threshold adjustment, so the English estimate may be biased downward; if it is, by roughly the 2.7-point shortfall seen in Mex-Cog, the English harmonised-protocol value would be about 9.7% and the under-detection ratio about 7.8 rather than 5.7 — a correction that strengthens rather than weakens the conclusion while making the 1.4-fold harmonised-protocol spread even narrower. The under-detection conclusion nonetheless holds whichever English value is used (ratios 4.96–10.4); the exact magnitude does not. One further data caveat: the ELSA age variable is about three years behind the wave-10 interview; re-computing the English estimate with the wave-10 age variable gives 1.07% rather than 1.25% and raises the English–European contrast to 4.9-fold (3.4–7.2), so the age handling makes our estimates conservative. Second, no published CHARLS-HCAP prevalence exists, so the harmonised-protocol comparison covers three of our four cohorts; the Chinese arm of the impossibility argument therefore rests on a national door-to-door survey rather than on HCAP, although the conclusion is robust across a grid of plausible true values. Third, CHARLS does not administer backward counting, so two points were imputed for all Chinese respondents; because the item carries almost no information anywhere (loadings 0.02–0.09) this is unlikely to affect the conclusions, but it does mean that the Chinese score is a three-item scale in practice. Fourth, the proxy branch in SHARE was calibrated rather than directly measured, and sensitivity analyses varying proxy prevalence by ±50% moved the SHARE estimate only between 5.04% and 5.48% (and extreme assumptions in which proxies carry the same or 2.2 times the self-report prevalence moved it to 4.88–4.95%), so this assumption does not drive the cross-national comparison. Fifth, equated prevalence values are model-based counterfactuals, not measurements, and the equating models fit only moderately well (CFI 0.884–0.924; RMSEA 0.097–0.185); we therefore report equating as one line of evidence among several rather than as the primary estimate. Sixth, we analyse prevalence cross-sectionally and cannot address incidence, and ELSA does not release death status in the relevant waves, so the mortality anchor rests on three cohorts. Seventh, differential item functioning and genuine population differences in the latent trait are not separately identified without external anchors; our partial-scalar solution provides a lower bound on the measurement contribution, not an exact decomposition. Eighth, misclassification risk in algorithmic classification is not random: it has been shown to vary systematically with race and ethnicity in the USA [29], and our own results show the same for education, so the country-level bias we document is likely to be heterogeneous within countries as well.

## Conclusion

Across four population-based cohorts, the same brief dementia algorithm yields prevalence estimates that differ tenfold, but the difference is not identifiable as disease: it depends on analytical choices, on a single poorly functioning item, and on operating characteristics that cannot be simultaneously true in the USA and China. Standardised clinical assessment narrows the same comparison to about 1.4-fold, and to no more than 1.9-fold under any available English estimate. Cross-national dementia prevalence should therefore be reported as a specification range or an equated interval, with an explicit invariance statement and an enrichment ratio — not as a single number. What remains more stable is the partial ordering of populations and, more clearly, the relative risk attached to the label; what does not is the count, and therefore the burden, on which policy currently relies.

## Ethics statement

Each cohort was approved by its own ethics body and all participants gave written informed consent: the Health and Retirement Study by the University of Michigan Institutional Review Board; the English Longitudinal Study of Ageing, including the ELSA-HCAP wave, by the UK National Research Ethics Service; the Survey of Health, Ageing and Retirement in Europe by the Ethics Council of the Max Planck Society together with national committees in the participating countries; and the China Health and Retirement Longitudinal Study by the Peking University Biomedical Ethics Committee. This study used de-identified data obtained through each cohort’s official data-access procedures and required no additional ethical approval.

## Author contributions

**Sumei Zhu:** Conceptualization, Data curation, Formal analysis, Investigation, Methodology, Software, Validation, Visualization, Writing – original draft. **Yukai Luo:** Conceptualization, Methodology, Project administration, Resources, Supervision, Writing – review & editing. Both authors read and approved the final manuscript and agree to be accountable for all aspects of the work. The authors received no specific funding for this work.

## Data availability

The HRS, ELSA, SHARE and CHARLS data are available to registered researchers from their respective distributors: the Health and Retirement Study (hrs.isr.umich.edu), the UK Data Service (SN 5050 for ELSA and SN 9081 for the ELSA-HCAP wave analysed here), SHARE (share-project.org) and CHARLS (charls.pku.edu.cn). All analysis scripts, derived datasets and figure-generating code are archived at Zenodo (DOI: 10.5281/zenodo.24166170), and the pipeline reproduces every table and figure from the released cohort files.

## Acknowledgments

The authors thank the HRS, ELSA, SHARE and CHARLS teams for data collection and distribution, and the participants of all four cohorts.

---

**S1 Fig. Educational inequality in dementia labels depends on the definition.** (A) Weighted prevalence by education group under the 27-point algorithm, a lenient ≤9 cut-off and self-reported physician diagnosis. (B) Relative index of inequality by definition and cohort on a logarithmic scale; under self-reported diagnosis the Chinese gradient reverses (low–high difference −6.4 points; concentration index +0.060), and China moves from first to last in the cross-national ranking.

**S2 Fig. Derivation of the analytic sample in each cohort.** Interviewed respondents aged ≥65, the subset with a classifiable cognitive assessment (self-report 27-point score or proxy module), and the split between self-report and proxy branches (proxy counts refer to proxies with a classifiable assessment: 715 of 721 in HRS and 443 of 1,347 in SHARE; Table 1 reports all proxy interviews). Frame-level counts are not shown because the released files do not include a codebook distinguishing non-interview response codes.

## Supporting information

All supporting information is supplied as one document (Supporting Information), which contains S1-S14 Tables, S1-S2 Figs and the STROBE checklist. Machine-readable copies of every table are available from the corresponding author.

**S1 Table.** Cohort definitions, variable mapping and analytic samples. **S2 Table.** Proxy-branch calibration and sensitivity analyses. **S3 Table.** Measurement invariance under alternative estimators and balanced group sizes. **S4 Table.** Equating model fit and linked prevalence. **S5 Table.** Latent class model selection and parameters. **S6 Table.** Full specification curve (1,152 specifications). **S7 Table.** Multiple imputation and delta adjustment. **S8 Table.** Mortality models. **S9 Table.** Bias-correction and impossibility grids. **S10 Table.** Harmonised Cognitive Assessment Protocol sources and data-access routes. **S11 Table.** Standardisation weights and standardised estimates. **S12 Table.** Educational and sex inequalities by definition and cohort. **S13 Table.** Backward-counting imputation sensitivity. **S14 Table.** Robustness of the cross-national range: contemporaneous comparisons, bounds under extreme assumptions about unclassified CHARLS respondents, and composition of the specification space.

**S1 Fig.** Educational gradient and relative index of inequality under three definitions of dementia; full legend in the Figure legends section. **S2 Fig.** Derivation of the analytic sample in each cohort; full legend in the Figure legends section. **STROBE checklist.** STROBE statement for cross-sectional studies, with the location of each item in this manuscript.

